# The Effect of COVID-19 on Distracted Driving: A Survey Study

**DOI:** 10.1101/2022.12.26.22283062

**Authors:** Ramina Javid, Eazaz Sadeghvaziri, Mansoureh Jeihani

## Abstract

The COVID-19 pandemic caused a significant shift in people’s travel behaviors and distractions while driving. This paper aims to investigate the impacts of the COVID-19 pandemic on distracted driving by comparing their behavior before and during the pandemic (from 3/1/2019 to 3/1/2021) in the state of Maryland using a stated preference online survey. Some 158 people were recruited for the survey. Participants were asked about their risky driving behaviors and self-reported distraction both before and during the pandemic. To analyze the results, the Chi-square and posthoc tests with the Bonferroni adjustment were applied. The results showed that during the pandemic, distraction dropped from 25% to 21%. The highest reported distracted driving behavior during the pandemic was using hands-free cell phones (64%), using GPS (75%), and eating or drinking (57%). The respondents’ daily trips have significantly decreased - about 44% below prepandemic rates. Moreover, using a binary logistic regression, it was revealed that the odds of becoming distracted among participants who used a handheld cell phone before and during the pandemic were 4.5 and 6.6 times higher than others, respectively. The findings of this study shed light on the causes of distraction before and during the pandemic.

## 1. Introduction

The COVID-19 pandemic’s impacts are being felt throughout all modes of transportation around the world, significantly altering human mobility patterns, and daily transportation-related behaviors (*1*). The U.S. Center for Disease Control and Prevention (CDC) recommended social distancing, quarantining, and working from home starting in March 2020. States and localities implemented these recommendations by closing schools and businesses, and residents were urged to stay at home. Travel demand fell across the board because of these unexpected and extraordinary shutdowns (*2–4*). Therefore, this pandemic caused a significant shift in people’s travel behaviors and patterns (*5*).

Although traffic has decreased dramatically since the outbreak of the COVID-19 pandemic, roads have become much riskier. According to the National Safety Council, 42,060 people died in car crashes in 2020, which is up 8% from 2019. Moreover, the number of crashes per million miles increased by 63% in the U.S. (*6, 7*). In addition, 4.8 million people were seriously injured in car crashes in 2020, raising questions about the prevalence of distracted driving (*5*). An analysis of 86,000 crashes that occurred on U.S. roads in 2020 showed that risky distracted driving habits like texting while driving, which is illegal in 41 states, were often implicated in the country’s record-breaking crash rates during the lockdown months (*8*). Experts, however, emphasized that distracted driving predates the pandemic and that the recent shift in travel habits merely highlights a long-standing problem. The National Highway Traffic Safety Administration (NHTSA) stated that 3,142 people died in distracted driving crashes in 2020 (*9*). Moreover, according to the Centers for Disease Control and Prevention (CDC), distracted driving kills more than eight people and injures more than 1,161 per day in the U.S. (*10*). The results of an online survey study related to distracted driving conducted in the U.S in 2020, showed that 41% of all drivers texted, 32% read emails, 29% read social media, and 36% accessed the internet while driving. Compared to 2010 or 2015, more drivers reported engaging in each task in 2020 (*11*). Although cell phones might help by supplying traffic data (*12, 13*), with phone distractions behind the wheel causing 57% of all crashes on U.S. highways, all stakeholders must work together to tackle the Distracted Driving epidemic (*8*). It is important to understand the impacts of the COVID-19 pandemic on daily travel behavior and distracted driving, as these changes in daily travel will subsequently affect the safety of drivers.

In March 2020, the first case of COVID-19 was confirmed in the state of Maryland, and initial statewide lockdowns took effect. Like any other state, this pandemic caused changes in travel behavior for Maryland residents. From 2015 to 2019, distracted driving caused more than 55,000 crashes per year on average. Moreover, in 2020 and during the COVID-19 pandemic, there were a total of 45,378 crashes caused by distracted driving in Maryland—a 20% decrease from the previous year (*14*). Vehicle miles traveled declined dramatically during the pandemic; however, it is uncertain how the rapid decrease in traffic congestion affected drivers’ tendency to engage in distracted driving. It is expected that there will be some impacts on road crashes and travel patterns due to the impacts of the pandemic since driving behavior and traffic patterns altered dramatically. Although many studies have been conducted on different aspects of the COVID-19 pandemic, to the best knowledge of the authors, no study has investigated the effects of the COVID-19 pandemic on distracted driving in the state of Maryland. Therefore, the objective of this study is to compare self-reported distracted driving and risky driving behaviors (using a cell phone while driving, eating, drinking, etc.) both before and during the pandemic. To reach this goal, a state preference survey was conducted in the state of Maryland.

## 2. Background

The avoidance of driver distraction has always been a priority for the traffic safety community. According to the National Highway Traffic Safety Administration (NHTSA), distractions can be caused by anything that takes a driver’s attention away from the task of safe driving, including general inattention (lost in thought), smoking, eating, grooming, reading, communicating with passengers, manipulating vehicle controls, and using electronic devices (*9*). There are several possible sources of in-vehicle technology distractions. Cell phone use and texting are the most alarming distractions (*15–19*). Studies have shown that cell phone use among experienced drivers (both dialing and talking) increases the probability of a crash by a factor of four (*20, 21*).

According to one study, using electronics or cell phones while driving is the most typical form of distraction for drivers of all ages. Drivers drive more slowly with greater speed variance and less attention on the road while they are using cell phones or other devices. Texting causes more lane changes and collisions (*22*).

### 2.1. Distracted Driving During the COVID-19 Pandemic

In 2020, the COVID-19 pandemic resulted in significant changes in traveler behavior. Early evidence suggests that people have shifted away from taking public transportation in favor of driving as a result of the pandemic (*23*). The pandemic has resulted in several state-mandated stay-at-home orders, yet distracted driving has made the roadways more dangerous than before.

Several studies have been conducted to investigate different aspects of road crashes due to the impact of the COVID-19 pandemic. According to the most recent National Safety Council (NSC) Distracted Driving Survey, 2% of drivers admitted to speeding due to less congested roadways (*5*). Another study (*24*) indicated that during the pandemic, major speeding almost tripled as a percentage of total speeding violations. Moreover, another study (20) indicated that during the pandemic, major speeding almost tripled as a percentage of total speeding violations. However, another study found that the shelter-in-place order resulted in approximately 15,000 fewer crashes per month and 6,000 fewer injuries or fatal incidents per month in the state of California (*25*). Preliminary data from the first 3/12020 also show a decrease in motor vehicle crashes (both fatal and non-fatal) in the U.S. (*26*). Similarly, the number of vehicle crashes decreased steadily as the COVID-19 infection rates rose, according to a review of collision data from Florida, New York, and Massachusetts (*27*).

The pandemic may have different impacts on different demographic groups. Because of the pandemic, teen drivers are more likely to be involved in a crash due to distractions. According to NHTSA, teens are the most at risk behind the wheel, and the empty roads during the pandemic give them a false sense of protection (*28*). Minorities may be at increased risk during the epidemic for various reasons, some of which are related to the pandemic’s direct health effects, and others to the pandemic response’s implications. The elderly, those with underlying health problems, those experiencing poverty, those who belong to minority racial and ethnic groups, and those who live in rural areas all experience worse health outcomes. These groups, particularly those without reliable access to a household vehicle or who are unable to drive, frequently rely on others for transportation assistance (*29–31*). However, another survey study showed that although respondents admitted to driving less regularly, they also said they did not necessarily drive while distracted or more aggressively (*32*).

### 2.2. Distracted Driving in the State of Maryland

Distracted driving is most often caused by texting or talking on the phone, according to previous studies (*22*). The use of a handheld phone while driving is prohibited by law in Maryland. Driving while typing, sending, or reading a text or electronic communication can also result in a ticket. A driver that causes serious injury or death while talking on a handheld cell phone or texting may receive a prison sentence of up to three years and a fine of up to $5,000 (*33*). As shown in **Table 1**, according to the Maryland Department of Transportation Motor Vehicle Administration’s Highway Safety Office (MHSO) (*27*), distracted driving causes an average of 25,672 injuries and 189 fatalities every year in Maryland.

**Table 1.**
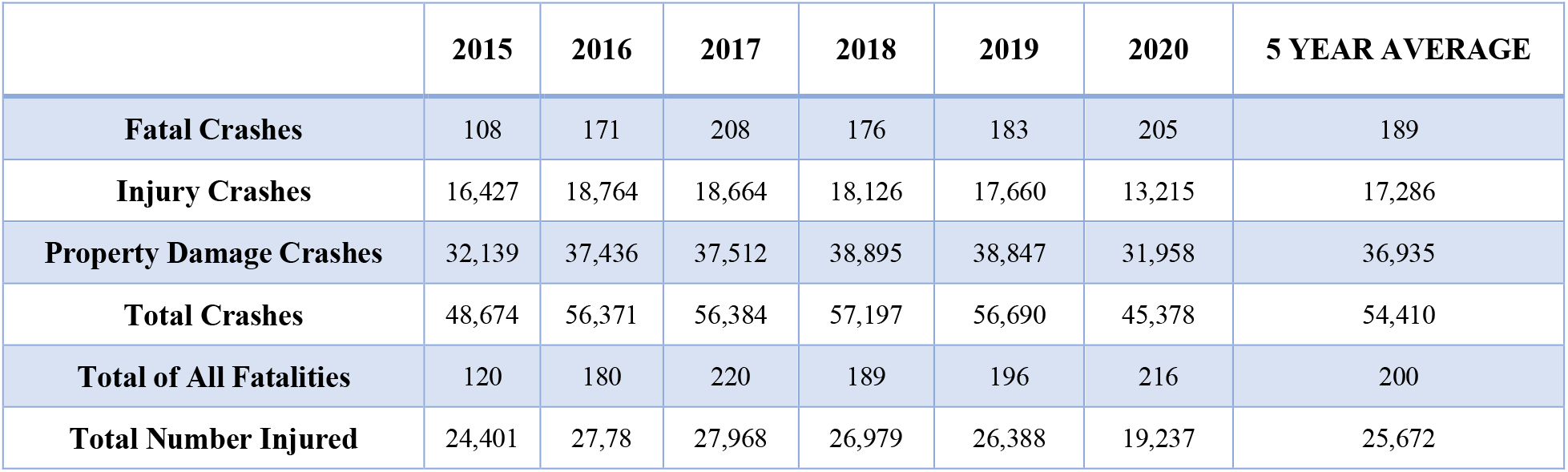
Distracted Driver Involved, 2015 – 2020 Crash Summary in Maryland

|  | 2015 | 2016 | 2017 | 2018 | 2019 | 2020 | 5 YEAR AVERAGE |
| --- | --- | --- | --- | --- | --- | --- | --- |
| **Fatal Crashes** | 108 | 171 | 208 | 176 | 183 | 205 | 189 |
| **Injury Crashes** | 16,427 | 18,764 | 18,664 | 18,126 | 17,660 | 13,215 | 17,286 |
| **Property Damage Crashes** | 32,139 | 37,436 | 37,512 | 38,895 | 38,847 | 31,958 | 36,935 |
| **Total Crashes** | 48,674 | 56,371 | 56,384 | 57,197 | 56,690 | 45,378 | 54,410 |
| **Total of All Fatalities** | 120 | 180 | 220 | 189 | 196 | 216 | 200 |
| **Total Number Injured** | 24,401 | 27,78 | 27,968 | 26,979 | 26,388 | 19,237 | 25,672 |

From the beginning of the pandemic (March 2020) until June 2022, there were 1,168,190 total confirmed COVID-19 cases and 14,912 deaths in the state of Maryland (*34*). **Fig. 1** shows the trend from January 2020 to March 2021 (this period was selected because of the survey questions). The period from December 2020 to January 2021 had the highest number of new cases per 1,000 people, after which the figure shows a downward trend.

**Fig. 1.**
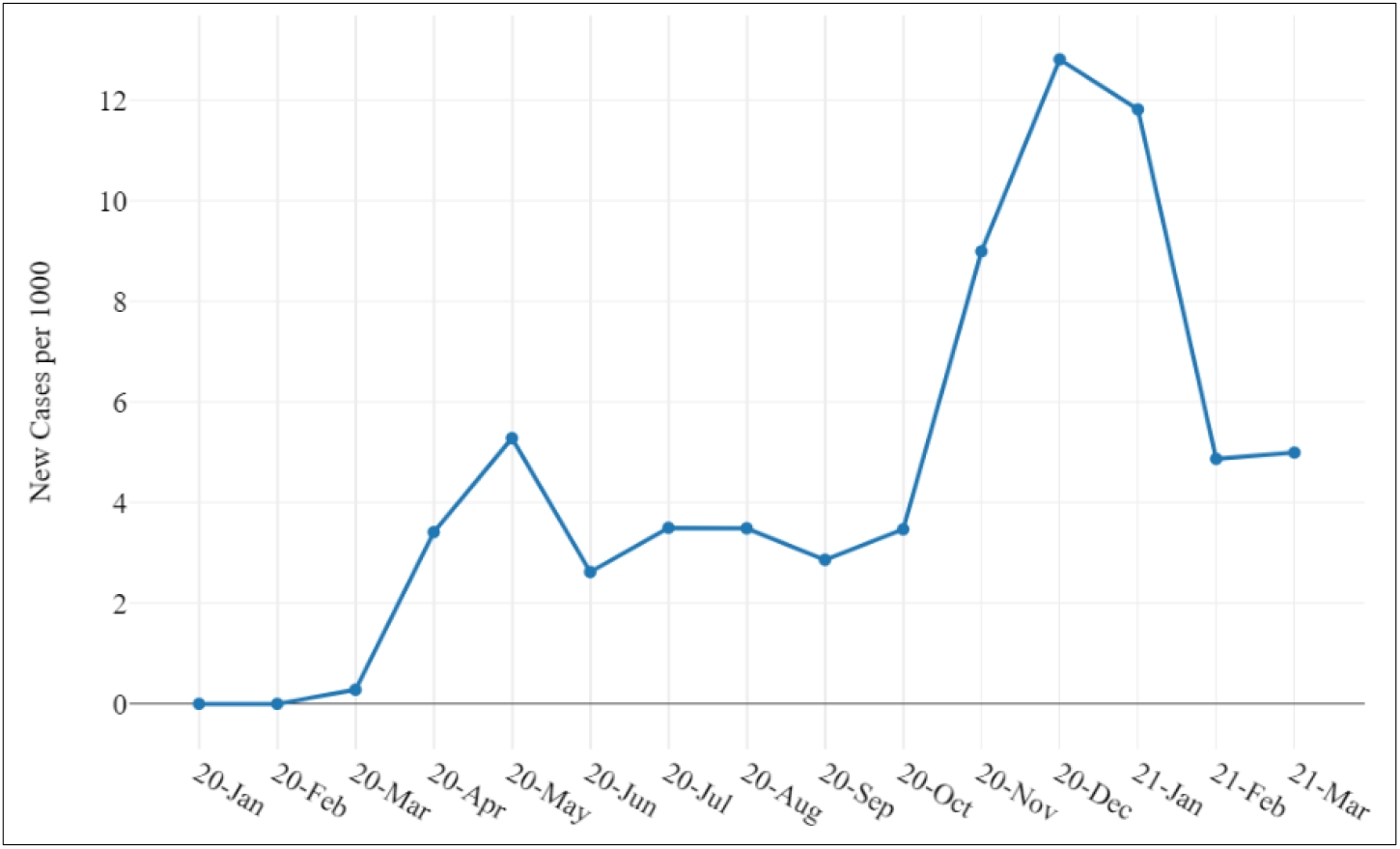
Number of COVID-19 daily new cases per 1,000 people in Maryland.

**Fig. 2** shows a dramatic decline in work trips per person per day from February to May 2020. After that, the number of work trips increased. Due to the outbreak of the COVID-19 pandemic earlier that year, starting in February, people had to stop working, or they worked remotely from home, which affected the driving behavior of workers (*35*).

**Fig. 2.**
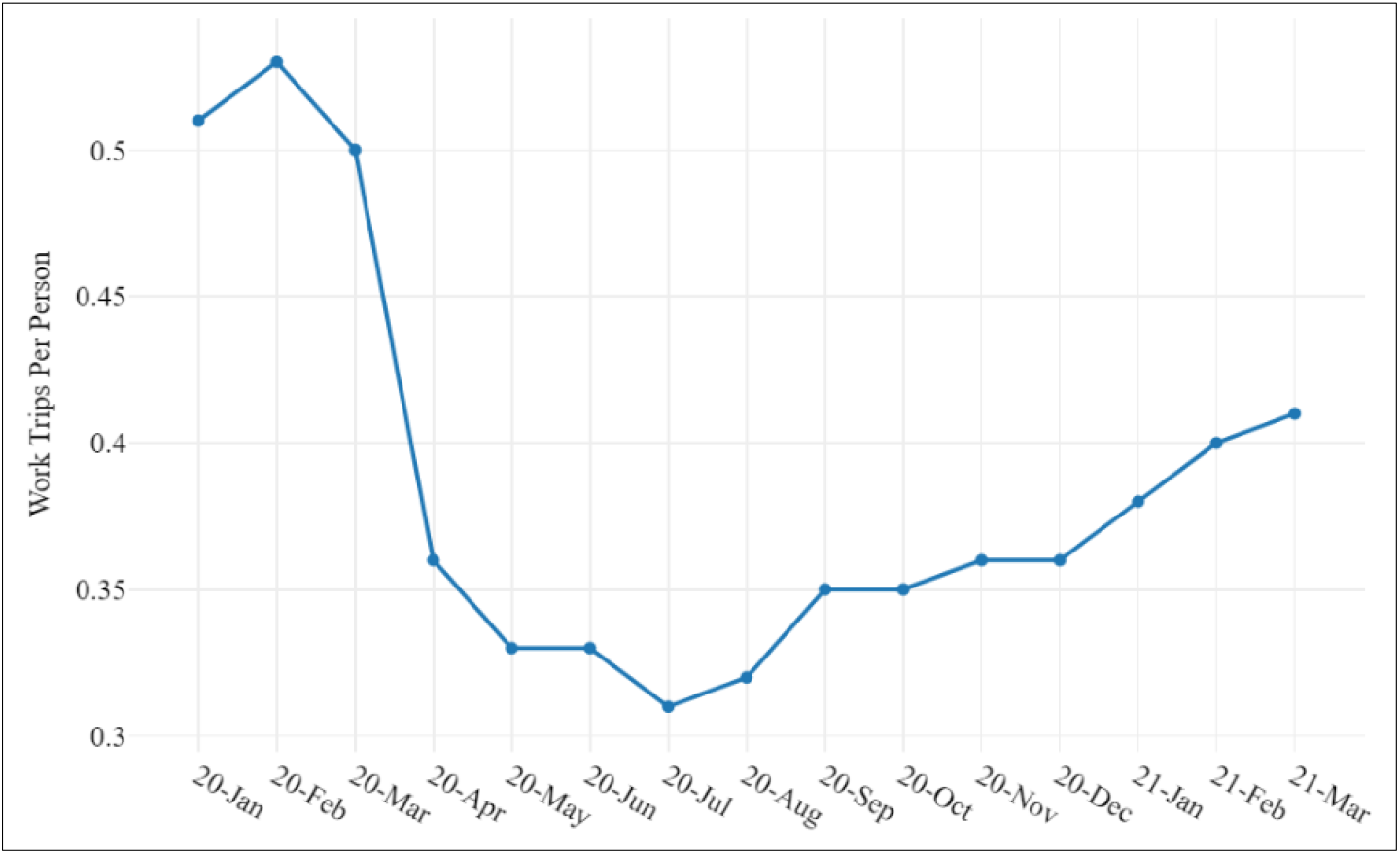
Number of work trips per person per day in Maryland.

As of June 15, 2021, the end of the COVID-19 pandemic state of emergency was announced (*34*). Events in 2020 related to the COVID-19 pandemic response were unprecedented. Due to the varying levels of statewide reactions and constraints, these shifting patterns and difficulties may change from state to state.

### 2.3. Summary of The Literature Review

The existing literature on the impact of COVID-19 on distracted driving indicates that the effects of the COVID-19 pandemic on travel patterns and driving behavior vary depending on regional characteristics. In conclusion, these preliminary data points to a surge in unsafe road user behavior, such as speeding and drunk driving, and reveal that the public safety measures implemented to combat COVID-19 resulted in lower road use (particularly during peak weekday travel times). Additionally, some preliminary data on crashes indicate a decline in both fatal and non-fatal crashes, while other data indicated a decline in overall crashes but an increase in more serious collisions.

Essentially, there is limited literature that has explored the COVID-19 pandemic’s impacts on distracted driving. The global outbreak of the pandemic brought many modes of transportation to a halt, with significant implications for all forms of transportation. The transportation networks and systems started to look very different during this pandemic. Existing research has made significant contributions to the methods and data analysis related to transportation during the COVID-19 pandemic; however, the effect of the COVID-19 pandemic on Maryland’s road safety is still largely unexplored, and the extensive influence of the COVID-19 pandemic on distracted driving and distracted driving behaviors still needs to be investigated. Therefore, this study aims to investigate self-reported driving behaviors using a stated preference survey and compare drivers’ behaviors before and during the pandemic. The following sections will present the methodology, results, and discussion of the study.

## 3. Data and Methodology

For this study, participants were recruited from Maryland State via manually distributed flyers, as well as online and through social media. Participants were required to be older than 16, have a valid driver’s license, and provide consent to participate in the research. Data were collected from a questionnaire-based survey using an online platform. Survey responses were monitored dynamically, and inattentive respondents were removed from the final data set by the authors. Finally, some 158 responses were finalized for analysis. The questionnaire took approximately 10 to 15 minutes to complete and was conducted between April 1, 2021, and May 1, 2021, via an online platform called Qualtrics.

### 3.1. Survey Design and Procedure

The questionnaire consists of three sections. The first section covered basic sociodemographic information about the participants (e.g., age, gender, education, employment, etc.). The second section consisted of self-reported risky driving behaviors (e.g., using hands-free or handheld cell phones, eating or drinking, etc.) before the pandemic (from March 1, 2019, to March 1, 2020). The third section consisted of the same questions however referred to the period during the pandemic (from March 1, 2020, to March 1, 2021). **Table 2** shows the questions asked in the survey.

**Table 2.** Summary of the Survey Questions

| Question |
| --- |
| What is the average annual driving mileage on your own car (in miles)? (Before the Pandemic, During the Pandemic) |
| How many miles do you drive per week on average? (Before the Pandemic, During the Pandemic) |
| Did you usually get distracted while driving? (Before the Pandemic, During the Pandemic) |
| Did you usually indulge in the following activities while driving Before the Pandemic (From 3/1/2019 to 3/1/2020)?<br>- Talk on the phone (hands-free)<br>- Talk on the phone (handheld)<br>- Texting<br>- Voice to text<br>- Read/update social media<br>- Read/respond to emails<br>- Take pictures/record video<br>- Using GPS<br>- Eat/drink<br>- Raking on/off clothes |
| Did you usually indulge in the following activities while driving During the Pandemic (From 3/1/2020 to 3/1/2021)?<br>- Talk on the phone (hands-free)<br>- Talk on the phone (handheld)<br>- Texting<br>- Voice to text<br>- Read/update social media<br>- Read/respond to emails<br>- Take pictures/record video<br>- Using GPS<br>- Eat/drink<br>- Raking on/off clothes |
| How many times have you experienced a near-crash experience due to using a cell phone while driving? |
| How many times have you experienced a crash due to distraction (such as using a cell phone or any kind of in-vehicle technology) while driving in the last two years? |

The composition of the participant samples and the socio-demographic information of the respondents is presented in **Table 3**.

**Table 3.** Socio-demographic Information of Participants

| Variables | Variable | Percent |
| --- | --- | --- |
| **Gender** | Female | 50.63 |
|  | Male | 49.36 |
| **Age** | 16 to 19 | 2.53 |
|  | 20 to 24 | 5.06 |
|  | 25 to 29 | 8.22 |
|  | 30 to 34 | 10.75 |
|  | 35 to 39 | 15.82 |
|  | 40 to 44 | 12.02 |
|  | 45 to 49 | 8.22 |
|  | 50 to 54 | 5.69 |
|  | 55 to 59 | 10.12 |
|  | 60 to 64 | 8.22 |
|  | 65 or more | 13.29 |
| **Education Status** | Less than high school graduate | 1.89 |
|  | High school graduate, including GED | 7.59 |
|  | Some college or associate degrees (e.g., AA, AS) | 18.35 |
|  | Bachelor's Degree (e.g., BA, AB, BS) | 27.21 |
|  | Graduate or professional degree (e.g., MA, MS, MED, Ph.D., MD, DDS) | 44.93 |
| **Employment Status** | Yes, Full-time | 58.86 |
|  | Yes, Part-time | 12.65 |
|  | No | 28.48 |

### 3.2. Data Analysis

Several variables were investigated to better understand the drivers’ behavior before and during the COVID-19 pandemic. Summaries of trends to understand and represent the variables before and during the COVID-19 pandemic are presented with graphs. Many studies use statistical models to develop policies to improve traffic safety, investigate and forecast travel behavior, and pinpoint deficiencies in transportation policy (*12, 13, 22, 36–39*).

In this study, the Chi-square and Fisher’s Exact tests are used to determine the association between variables. If the association between two variables was significant, Cramer’s V was used to determine the strengths of the association. Moreover, contingency tables (also known as crosstabulation or crosstab) were used to show the (multivariate) frequency distribution of the variables to find interactions between them. Also, cell residuals, including standardized residuals and adjusted residuals, are used in testing for cell significance, which is known as a post-hoc test, after a statistically significant Chi-square test. For Post Hoc tests following a Chi-square, we use the Bonferroni adjustment. This adjustment is used to avoid a type I error when multiple comparisons are made (*40, 41*). Moreover, to test whether the changes before and after the pandemic are statistically and significantly different in proportion, McNemar’s test was used. All statistical analyses are conducted with a 95% level of confidence. RStudio was used for data processing and analysis (*42*).

## 4. Results

The results of the survey questions are presented in this section. The frequency and average mileage of driving, the relationship between the participants’ demographic information and their self-reported distracted driving behavior before and during the pandemic, and incidents of risky driving behavior, near-crashes, or crashes due to distraction were analyzed in this section.

### 4.1. Frequency and Average Mileage of Driving

The frequency of driving respondents reported before and during the COVID-19 pandemic shows that before the pandemic, most participants drove every day (55%); however, after the pandemic, this figure dropped sharply (11%). **Fig. 3 (a)** shows that during the pandemic, about 68% of participants were driving less than 8,000 miles annually. Also, the proportion of travelers driving 30,000 miles or more annually dropped to less than 3% during the pandemic. **Fig. 3 (b)** shows the weekly driving mileage of participants. Most of the participants, about 70%, drove less than 100 miles weekly during the pandemic.

**Fig. 3.**
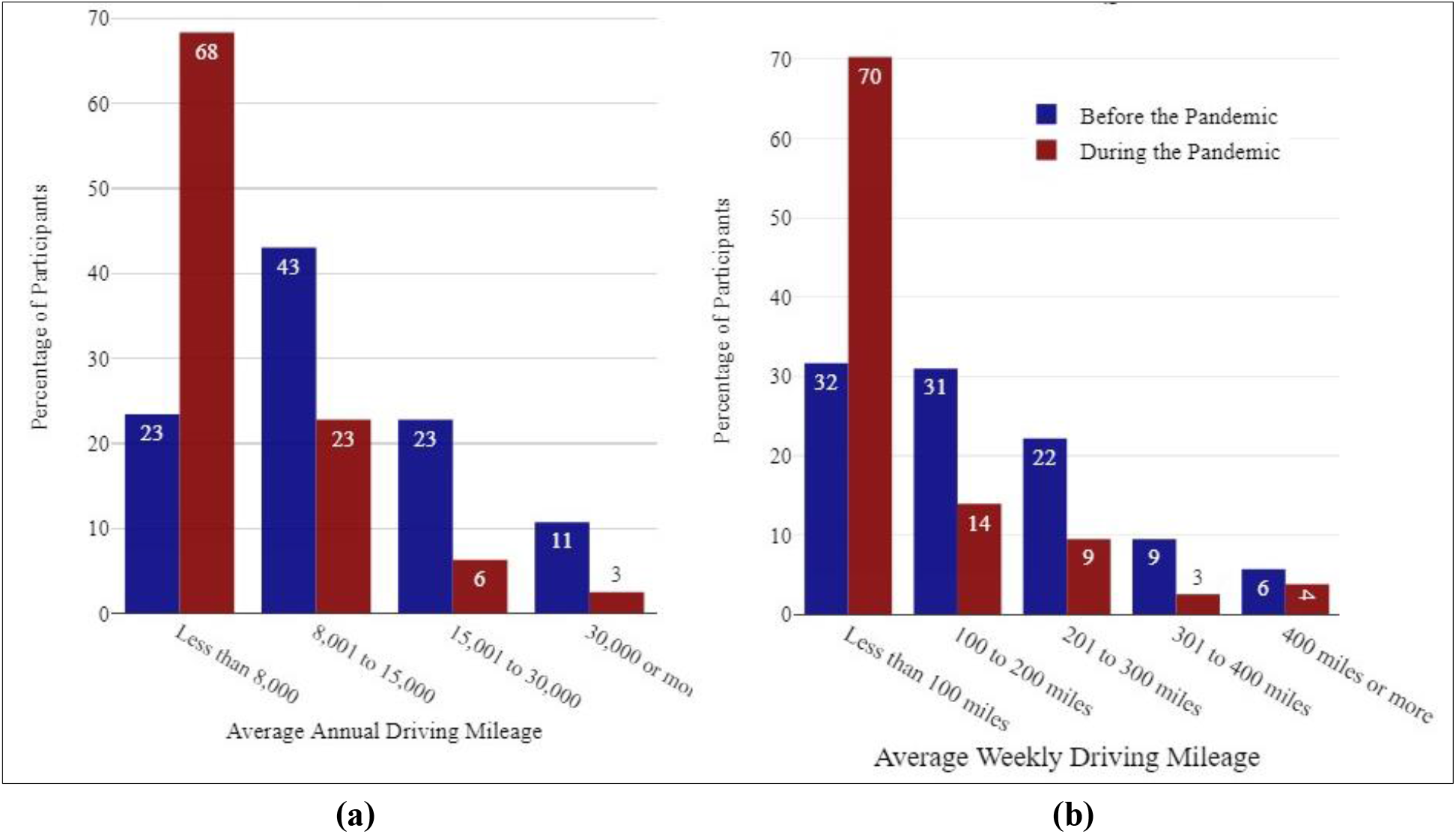
(a) average annual driving, and (b) average weekly driving.

### 4.2. Self-reported Distraction Driving

The self-reported distracted driving rates decreased by 4% during the pandemic, from 25.3% to 21.5%. A Chi-square Bonferroni Post Hoc test (p-value<0.0001) revealed that self-reported distraction dropped significantly before and during the pandemic. However, the Chi-square test revealed no association between getting distracted before and during the COVID-19 pandemic and the different socio-demographic information of drivers, including gender, age, income groups, levels of education, or race.

### 4.3. Risky Driving Behaviors

The findings show that hands-free cell phone use while driving dropped by 5% (from 68% to 63% of users) during the COVID-19 pandemic (**Fig. 4**). The p-value of the Chi-square test (0.02686) shows that the changes in the proportion of using a hands-free cell phone before and during the pandemic are statistically significant. Handheld cell phone use remained unchanged at 16% before and during the COVID-19 pandemic. Texting (20%) and using voice-to-text (24%) while driving did not change significantly before and during the pandemic. Other activities that include using a cell phone while driving is reading or updating social media (13%), reading or responding to emails (13%), and taking pictures or recording videos (12%), none of which changed significantly before or during the pandemic.

**Fig. 4.**
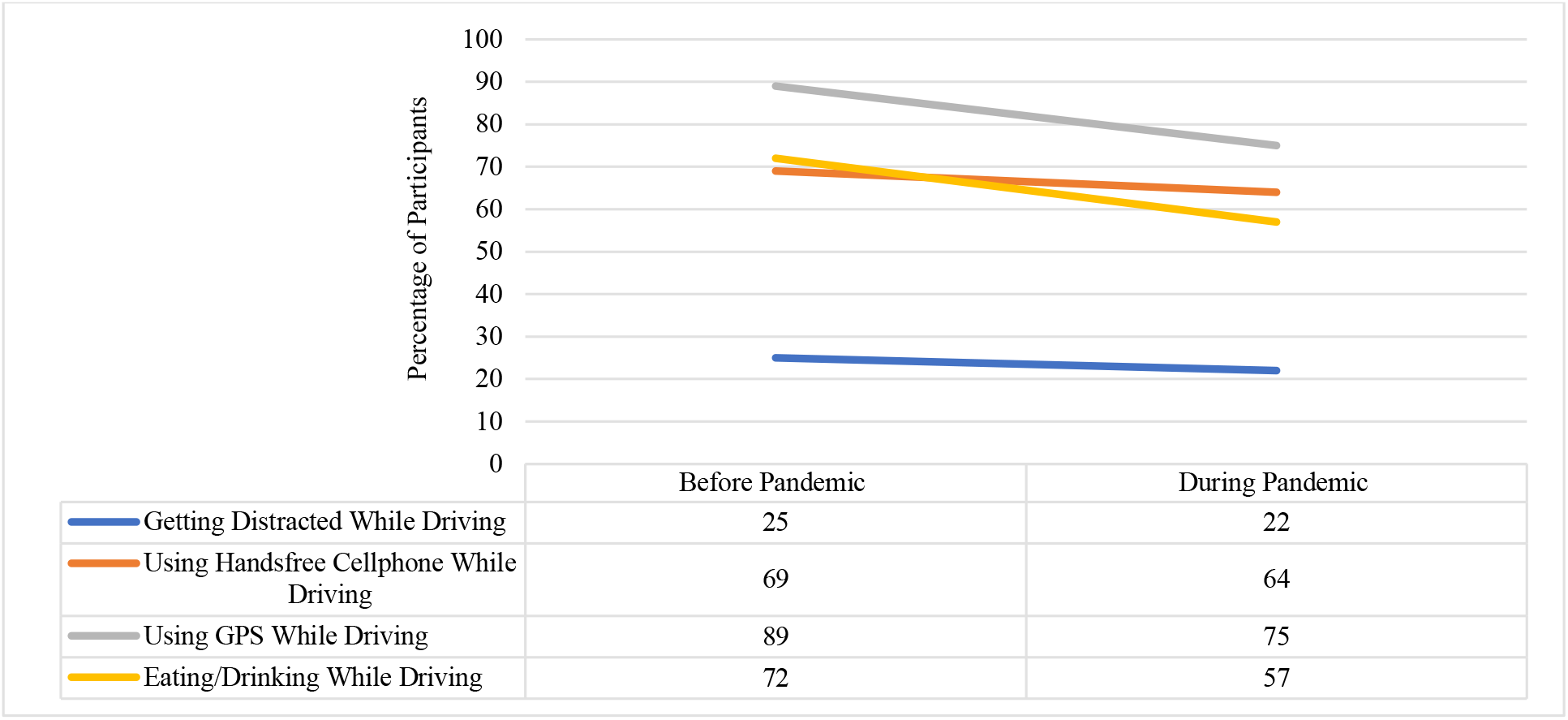
Different distractions before and during the pandemic.

Other distractions include using GPS, eating or drinking, and taking off or putting on clothes. Taking off or putting on clothes while driving did not change significantly before and during the COVID-19 pandemic (90%). However, the findings clearly show that using GPS while driving dropped 14% before (89%) and during the pandemic (75%). The p-value of the Chi-square test (7.562e-06) indicates that the changes in the proportion of using GPS before and during the COVID-19 pandemic are statistically significant. Eating or drinking while driving dropped 15% before (72%) and during the pandemic (57%). The p-value shows a statistically significant change in the proportion of eating or drinking before and during the COVID-19 pandemic (p-value of the Chi-square test = 1.383e-05).

### 4.4. Crash and Near-crash Due to Distraction

According to the survey’s results, almost 12% of participants had at least one crash due to distraction in the past two years (2019-2021). A Chi-square Bonferroni Post Hoc test (p-value<0.0001) revealed that the likelihood of having at least one crash due to distraction dropped significantly (23%) between 2019 (before the COVID-19 pandemic) and 2020 (during the COVID-19 pandemic). All the crashes fit in “property damage only” crashes. Moreover, the most frequent types of crashes were rear-ended and U-turn.

Similarly, the likelihood of having at least one near-crash experience dropped 19% from before the COVID-19 pandemic (31%) and throughout the pandemic (12%). Not having a near-crash experience increased by 30% during the pandemic (as shown in **Fig. 5**). A Chi-Square Bonferroni Post Hoc test (p-value<0.0001) revealed there is a significant difference between not having a near-crash experience due to distraction before and during the COVID-19 pandemic.

**Fig. 5.**
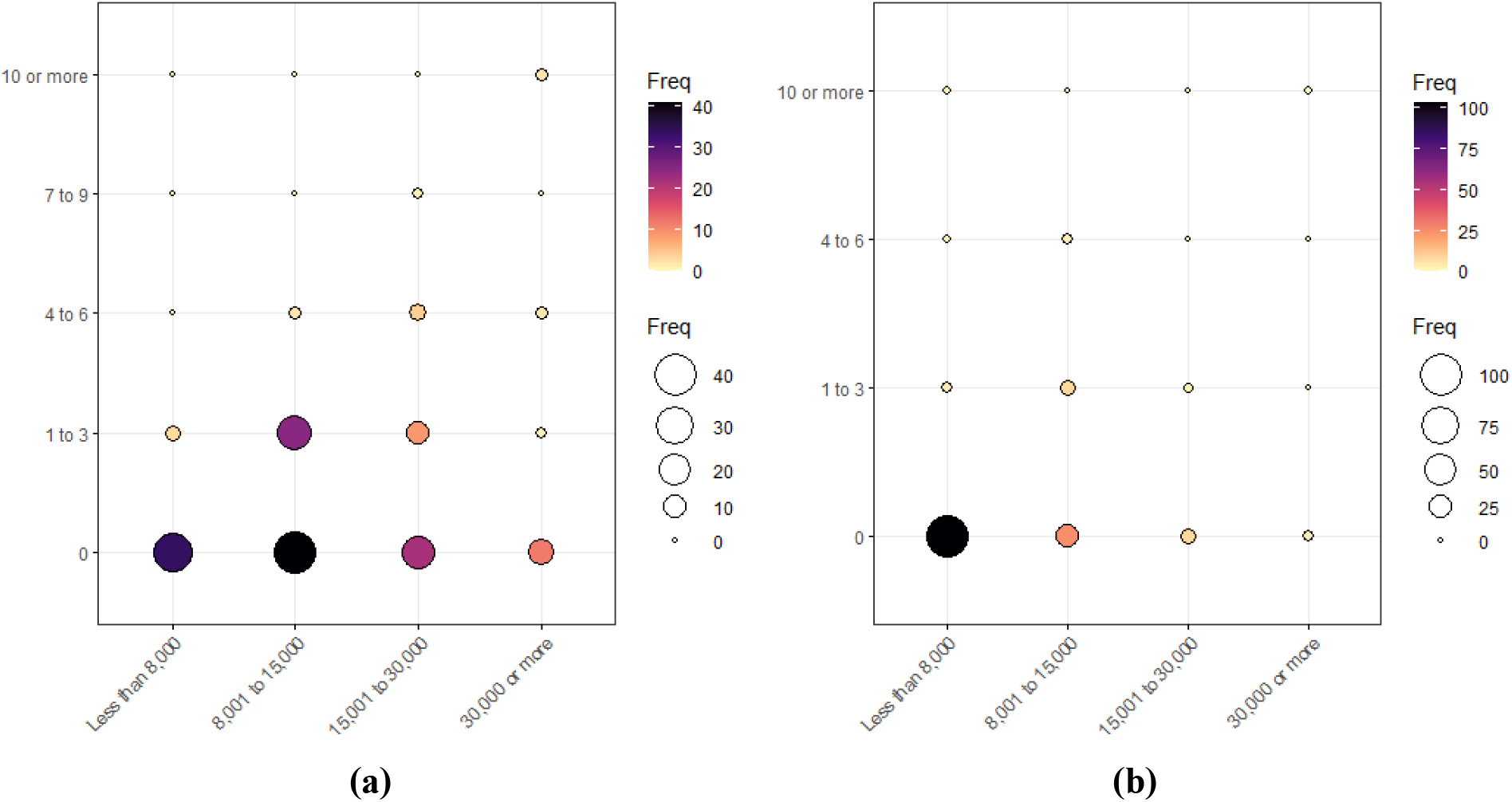
(a) Number of near-crash experiences while driving in one year versus average annual driving before the pandemic, (b) Number of near-crash experiences while driving in one year versus average annual driving during the pandemic.

An analysis using Pearson’s Chi-squared test and a Chi-square Bonferroni Post Hoc test revealed that a statistically significant relationship exists for changes before and during the COVID-19 pandemic regarding distracted driving behavior in Maryland. Average annual/weekly driving mileage, hands-free cell phone use, GPS usage, eating or drinking while driving, and not having a crash or near-crash incident due to distraction all significantly decreased during the pandemic (from March 1, 2020, to March 1, 2021).

### 4.5. Distracted Driving Behavior Model

A binary logistic regression was developed to predict the odds of getting distracted based on the values of the independent variables. **Table 4** presents the results of the binary regression model before and during the pandemic. Before the pandemic, using a hands-free or a handheld cell phone was significant, meaning that the odds of being distracted among the participants who used a hands-free cell phone is exp(1.44) = 4.2 times higher than others. Moreover, the odds of being distracted among the participants who used a handheld cell phone is exp(1.5) = 4.5 times higher than others before the pandemic. The odds of being distracted among the participants who use a handheld cell phone increased during the pandemic, to exp(1.8) = 6.6 times higher than others during the pandemic. Since the number of trips decreased during the pandemic, GPS use decreased as well. The odds of being distracted by a GPS reduced dramatically during the pandemic. Although the difference between using GPS before and during the pandemic was not significant, the odds of being distracted among the participants who used GPS before the pandemic were higher than others. However, the odds of getting distracted among participants who used GPS during the pandemic were lower than for those who did not.

**Table 4.** Results of Binary Logistic Regression Model Before and During the Pandemic

| Variable | Before the Pandemic |  |  | During the Pandemic |  |  |
| --- | --- | --- | --- | --- | --- | --- |
|  | Estimate | Pr(>\|z\|) | Code | Estimate | Pr(>\|z\|) | Code |
| (Intercept) | -21.04 | 0.99 |  | -3.06 | 0.00 | ** |
| Hands-free | 1.44 | 0.04 | * | 0.89 | 0.17 |  |
| Handheld | 1.50 | 0.01 | * | 1.88 | 0.00 | ** |
| Texting | 0.63 | 0.31 |  | 0.48 | 0.47 |  |
| Voice to Text | 0.56 | 0.25 |  | 0.95 | 0.08 | . |
| Social Media | 0.73 | 0.34 |  | 1.51 | 0.06 | . |
| Emailing | 0.03 | 0.97 |  | -1.09 | 0.20 |  |
| Taking Picture | 0.06 | 0.93 |  | -0.19 | 0.82 |  |
| Using GPS | 17.38 | 0.99 |  | -0.39 | 0.70 |  |
| Eat or Drink | 1.08 | 0.13 |  | 0.74 | 0.31 |  |
| Taking on / off Clothes | -1.46 | 0.11 |  | -0.86 | 0.35 |  |
| Null deviance: 178.79 on 157 degrees of freedom |  |  |  | Null deviance: 164.56 on 157 degrees of freedom |  |  |
| Residual deviance: 131.48 on 147 degrees of freedom |  |  |  | Residual deviance: 124.55 on 147 degrees of freedom |  |  |
| AIC: 153.48, R2 (Nagelkerke): 0.3819047 |  |  |  | AIC: 146.55, R2 (Nagelkerke): 0.3456938 |  |  |
| Signif. codes: 0 '***' 0.001 '**' 0.01 '*' 0.05 '.' 0.1 ' ' 1 |  |  |  |  |  |  |

## 5. Discussion

The current study aims to investigate changes in driving behavior and distracted driving behavior before and during the COVID-19 pandemic. Some 158 participants were recruited to fill out a stated preference survey in the state of Maryland. The results suggested that the statewide distracted driving rate decrease was influenced by a decline in cell phone use while driving (handsfree) and a decrease in non-cell phone-related distractions (eating or drinking). A similar pattern was observed in the reduction of crashes due to distraction and a decrease in average annual and weekly driving, which is in line with previous studies related to COVID-19 and distracted driving. Distracted driving predates the pandemic, and the recent shift in travel habits merely highlights a long-standing problem. The governor’s shelter-in-place order and similar orders in other jurisdictions profoundly affected daily travel in Maryland. According to the results of this study, everyday trips significantly decreased to about 44% below pre-pandemic rates in Maryland, and the number of those driving 30,000 miles or more dropped significantly. This may be because daily commutes, work-related long-distance trips, and non-work travels decreased during the pandemic.

Moreover, there was a significant association between getting distracted before and during the pandemic, with distraction rates dropping about 4% among respondents. According to the results, no significant relationship was found between the demographics or characteristics of drivers and self-reported distraction before and during the COVID-19 pandemic. We found that the number of people reporting at least one near-crash experience dropped by 19%, and not having a near-crash experience increased by 20% in Maryland. A possible explanation could be due to a significant decrease in traffic and frequency of driving statewide. A comparison of the results of this study with similar studies regarding distracted driving and the COVID-19 pandemic shows that the consequences of the pandemic differ according to geographic and demographic characteristics and may differ from state to state. As we progressed through the COVID-19 pandemic, transportation networks and systems began to take on new forms. Reduced car crashes and fatalities would be a silver lining of the COVID-19 pandemic. After all, it seems that more individuals working or studying from home will result in fewer cars on the road, lowering the risk of crashes. However, mental health difficulties have been on the rise during the pandemic. These issues influence every area of day-to-day living, including driving behaviors. Nevertheless, the extent of the COVID-19 pandemic’s impact on distracted driving has yet to be investigated in the long term. The shifting trends mentioned in this research will need to be followed to see if they are temporary or permanent, as well as how planning would need to be altered to accommodate current trends.

There are some limitations to this study. First, because the participants were asked to report their distracted driving behavior, it may somewhat affect their judgment. Second, it is worth mentioning that the sample size was limited to just over 150 respondents, which is, however, consistent with the sample sizes adopted in related studies (*32, 43, 44*).

## 6. Summary and Conclusion

This paper is a reliable approximation of the COVID-19 pandemic’s impacts on distracted driving in Maryland. This goal was reached by using an online questionnaire to measure and compare changes in distracted driving behavior before and during the COVID-19 pandemic in the state. This study used the Chi-square and Fisher’s test, Cramer’s V test, McNemar’s test, contingency tables, and a binary regression model to show the frequency distribution of the variables to find interactions between variables before and during the COVID-19 pandemic. Moreover, a Post Hoc test following a Chi-squared test with a Bonferroni adjustment is used for multiple comparisons.

Frequency of driving, average annual and weekly driving mileage, cell phone-related activities while driving (hands-free, handheld, texting, voice-to-text, reading or updating social media, reading or responding to emails, and taking a picture or recording a video), other distracted driving behavior (eating or drinking, using GPS, taking on or off clothes) and crash and near crashes due to distraction were investigated before (from March 1, 2019, to March 1, 2020) and during (from March 1, 2020, to March 1, 2021) the COVID-19 pandemic. Findings from the state-specific survey of 158 Marylanders from this study show different impacts across different distracted driving behaviors before and during the pandemic. Many people started driving much less frequently during the pandemic, opting to stay home instead. Everyday trips in Maryland decreased by roughly 44% during the COVID-19 pandemic, compared to prepandemic levels. Self-reported distractions dropped by 4% during the COVID-19 pandemic. Using GPS while driving was identified as the most common distracted driving behavior by 75% of respondents, followed by eating or drinking at 57%. Hands-free cell phone usage decreased by 5%, eating or drinking by 15%, and the use of GPS fell by 14%. Furthermore, using a binary logistic regression, it was discovered that participants who used a hands-free cell phone before and during the pandemic were 4.5 and 6.6 times more likely to become distracted than others.

Although existing research has assisted in the development of data analysis during the COVID-19 pandemic, additional research should be conducted to address some of the study’s limitations and further investigate the patterns discovered in this study. Behavioral surveys and additional analysis are urgently needed to corroborate some of the patterns suggested in this study about the COVID-19 pandemic and distracted driving behavior. Continual examination of the research’s results and trends is necessary to discover if these outcomes are temporary or permanent, as well as how planning will need to change to meet current trends.

## Data Availability

All data produced in the present study are available upon reasonable request to the authors

## Acknowledgments and Declarations

This study was financially supported by the Maryland Department of Transportation - Motor Vehicle Administration - Maryland Highway Safety Office.

All authors reviewed the results and approved the final version of the manuscript.

The authors declare that the contents of this article have not been published previously. All the authors have contributed to the work described, read, and approved the contents for publication in this journal. All the authors have no conflict of interest with the funding entity or any organization mentioned in this article that may have influenced the conduct of this research and the findings.

